# Comparative Performance of Machine Learning Models in Predicting Fertility Based on Insights from BDHS Data

**DOI:** 10.64898/2025.12.16.25342342

**Authors:** Md. Yusuf Hossain Ador, Samrat Kumar Dev Sharma, Md. Rukonuzzaman, Futanta Chakma, Md. Kamruzzaman

## Abstract

**Background:** Fertility is a social indicator that represents the country’s growth and economic sustainability. The fertility rate in a country signifies the average number of kids that a woman gives birth to throughout her lifetime. The current research is going to use several machine learning models in such a way that they would be capable of detecting the factors that are driving and are responsible for the fertility rate in Bangladesh.

**Methods:** The data used for this study was obtained from the Bangladesh Demographic Health Survey (BDHS), which was conducted in 2021-22. A variety of machine learning (ML) models and techniques were put into practice including Random Forests (RF), Decision Trees (DT), K-Nearest Neighbors (KNN), Logistic Regression (LR), Support Vector Machines (SVM), XGBoost, LightGBM, Neural Networks (NN), Stacking, and Voting. Along with K-fold cross-validation, Metrics of Accuracy, F1-Score (weighted), Precision (weighted), Recall (weighted), Area under the Receiver Operating Characteristics Curve (AUROC) (weighted), and the weighted average of the Confusion Matrix were applied to the assessment and comparison of the performance of the predictive models.

**Results:** This research unveils the discussion on traditional methods and Machine Learning methods, and we found that division, place of residence, religion, and wealth index), mother’s education father’s education father’s occupation), mother’s occupation, type of toilet facilities), and source of drinking water, contraception use were strongly associated with fertility. With the help of our best identified model Stacking, Voting, and Logistic Regression showed the best results with the highest accuracy (81%), F1-score (∼78%), and AUC ROC (81%), indicating strong and balanced predictive performance for predicting the determinants influencing the fertility in Bangladesh.

**Conclusion & Recommendations:** According to our study Stacking, Voting and Logistics Regression showed better prediction for predicting fertility in Bangladesh. Comparative with analysis with advanced techniques can be done in future work. Moreover, our policy makers and government can take necessary steps by focusing on key determinants influencing fertility.

## Introduction

Fertility is a social indicator that represents the country’s growth and economic sustainability. It is a significant demographic indicator for public policy, social welfare, economic growth, and population dynamics [1]. The fertility conduct of the country influences its size, composition, and structure. As of 2014, the Bangladesh Bureau of Statistics (BBS) approximated that population of Bangladesh was around 0.158 billion, with a population density of 1,070 people per square kilometer. During the period of 2011-2014, the population increase was 8 million, which means an annual increase of over 2.0 million people [2]. The Total Fertility Rate (TFR) has undergone a considerable reduction as per the Bangladesh Demographic and Health Survey (BDHS), 2014 [3]. The adoption of effective contraceptives was the primary cause of a decrease in fertility among Asian women from low-income families [4]. According to a study, the income index, healthy living, and other social elements that are required for a drop in reproduction have improved [2]. One of the main causes of the decreased fertility trend was the family planning program [5]. The CPR increased from 8% in 1975 to 44% in 1993–94 as a result of these initiatives. It can be concluded that the declining trend in fertility is not primarily caused by socioeconomic factors like female employment and education [6]. Human fertility is fundamentally a biological process, but it is also a community and individual issue due to the significant effect of society and cultural variables. Because a society’s ability to reproduce is essential to its survival, all societies have aspects of their cultures that support and encourage some degree of reproduction [7]. In some study. Muslims were found to have a greater TFR (2.6 versus 2.4) than non-Muslims. In terms of media access, we found that people without media exposure had a higher TFR (2.8) than people with media exposure (2.4) [8]. In terms of lowering fertility, different regions demonstrated different levels of success. The study’s main goals are to identify spatial dependence and look into the factors that have contributed significantly to the reduction in fertility. Globally, there are significant variations in rural-urban fertility [9]. In a comparable pattern, 41.9 percent of rural women had more than two children, compared to 33.2 percent of urban women. In Bangladesh, however, the disparity in fertility between rural and urban areas is significantly decreasing [10]. However, changes in background factors that affect reproduction, such media exposure, education, and money, may not always result in a change in fertility. In the real world, socioeconomic factors have an indirect effect on reproduction because of influencing proximate determinants [10]. Prior studies have demonstrated that a number of socioeconomic status and demographic variables are significant drivers of Bangladesh’s demand for fertility [11&12]. According to the majority of earlier studies, gender preference is the most important major factor influencing fertility preference in Bangladesh [12, 13]. Despite of many improvements, Bangladesh’s fertility rate is still high when compared to too many other developing nations [27], indicating the need for more research into the factors that influence fertility behavior.

In some previous study due to the direct effect between fertility and proximate determinants, Bongaarts aggregate fertility model was applied [14 & 26]. Furthermore, a study contributed to basic knowledge of the structural determinants of fertility rate in the Bangladesh by analyzing the expanded set of determinants at district level. The analysis extends the spatial model by allowing spatial dependence to vary across divisions and regions [15]. Traditional econometric models such as logistic regression [29] and probit models have been widely used to analyze fertility determinants [10, 12]. Even while these models are still quite useful, they have major drawbacks like detecting linear relationships and hypothesis testing, these models may fall short in capturing complex, nonlinear interactions among multiple variables [13].

Although numerous research has been done previously for discovering determinants and prediction in various sectors, there have been few studies to find fertility determinants and forecast using a machine learning approach. The principal aim of this research is to investigate the significant factors, measured through the number of children ever born, that correlate with fertility levels among females in Bangladesh. The study recognizes the importance of demographic, socio-economic, and health-related determinants influencing fertility behavior and thereby seeks to establish them as major contributing factors. Besides, the research intends to use ten different machine learning models for the purpose of predicting fertility levels, thus enabling data-driven approaches to be used for a more precise and insightful analysis. Ultimately, the study, using proper metrics, aspires to assess and compare the predictive capabilities of the different machine learning algorithms and thus pinpoint the best models for fertility prediction in the context of Bangladesh.

## Methodology

### Data source and study sample

This analysis was based on data taken from the 2022 Bangladesh Demographic and Health Survey (BDHS 2022), which is part of the internationally known Demographic and Health Surveys (DHS) Program [16]. The survey was implemented by the National Institute of Population Research and Training (NIPORT), with technical support from ICF International and financial assistance from international development partners. High-quality, nationally representative data on a variety of population, health, and nutrition variables in Bangladesh is provided by BDHS [16 & 17]. A stratified, two-stage cluster sampling design was employed to ensure representativeness across all eight administrative divisions, as well as urban and rural areas. In the first stage, 675 enumeration areas (EAs) were selected using probability proportional to size. In the second stage, a fixed number of households were systematically selected within each EA. All women aged 15–49 years residing in the selected households were eligible and invited to participate. A total of 30,078 women completed the interviews using the DHS Women’s Questionnaire [17]. These face-to-face interviews, conducted by trained field staff using Computer-Assisted Personal Interviewing (CAPI) devices, gathered detailed information on a variety of topics such as fertility and reproductive history, maternal and child health, nutritional status, contraceptive use, education, employment, household conditions, and access to health services. The entire data collection and processing workflow is illustrated in Fig 1.

**Fig 1:**
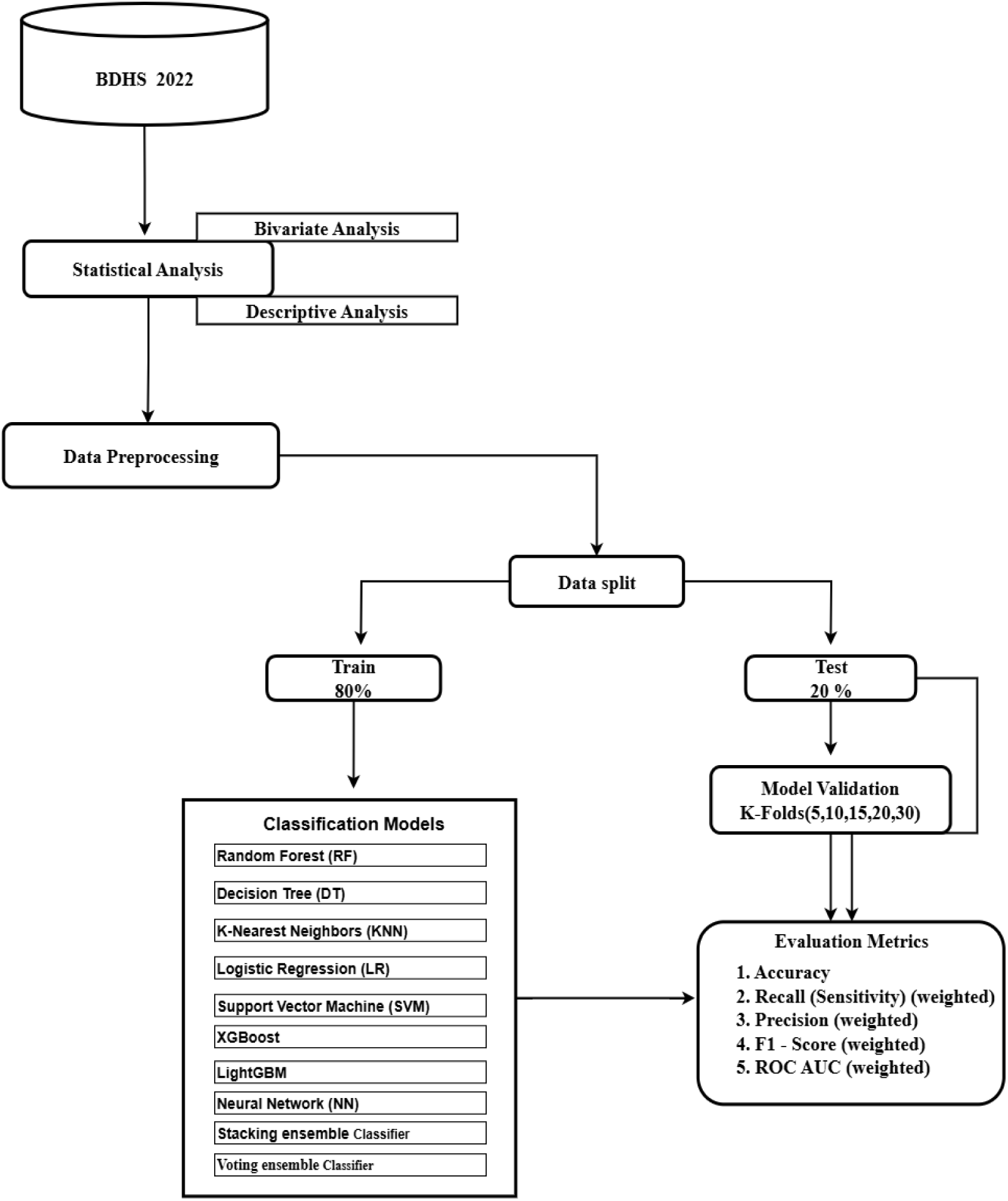
Overall workflow

### Study variables

#### Outcome Variable

The key end variable for this study was the number of children ever born (CEB) to women aged 15 to 49 years, which was divided into three categories: no children, one to three children, and four or more children. This classification reflects Bangladesh’s fertility patterns and enables for analysis of low, medium, and high fertility groups.

#### Predictors Variables

A total set of explanatory variables was chosen from the existing literature and their relation to fertility behavior. They included socio-demographic, economic, reproductive, and household-level variables [1, 2, 4, and 18]. Geographic location was determined by administrative divisions (Barishal, Chattogram, Dhaka, Khulna, Mymensingh, Rajshahi, Rangpur, and Sylhet) and type of residence (urban/rural). Religious affiliation was recorded as Muslim or non-Muslim. Socioeconomic status was quantified using the wealth index calculated by the DHS and divided into quintiles from poorest to wealthiest. Education and employment status of both women and partners were considered, categorized into conventional categories (no education, primary, secondary, tertiary) and types of employment (unemployed, agriculture, manual, professional, business, several others). Household-level environmental factors were toilet type, water source, and household size (1-3, 4-6, or >6 individuals). Nutritional status was measured using body mass index (BMI) and classified as underweight, normal weight, overweight, or obese. Reproductive and behavioral characteristics included age at first marriage (<18 or ≥18), contraception use (modern, traditional, none), child death experience (yes/no), media exposure (yes/no), and gender of household head (male/female). The woman’s ideal family size (0-2, 3-4, >4), the husband’s ideal (same, wants more, wants fewer, don’t know), and the father’s ideal family size (one to four) all indicated their marital status and fertility goals. These were employed to examine relationships with children ever born.

#### Descriptive and Bivariate Analysis

Descriptive statistical analysis was conducted to summarize the distribution of the study sample across all categorical variables. Frequency counts and percentages were calculated to describe the socio-demographic, economic, and reproductive characteristics of the women included in the study. To examine the bivariate associations between the number of children ever born (categorized as no children, 1 to 3 children, and 4 or more children) and the explanatory variables, Pearson’s Chi-square (χ²) test was employed [19]. This test assessed whether there were statistically significant associations between the outcome variable and each of the independent variables. All variables were included in the bivariate analysis regardless of their expected direction or strength of association. The results of the Chi-square tests are presented in **Table 1**. Statistical significance was determined using p < 0.05, with a focus on factors with highly significant connections (p < 0.001). These studies laid the groundwork for identifying potential factors for additional multivariable modeling.

**Table 1:** Descriptive analysis of demographic and background characteristics.

| Characteristics | Children Ever Born |  |  | Chi Square | P-value |
| --- | --- | --- | --- | --- | --- |
|  | No Children | 1 to 3 Children | 4 or more Children |  |  |
| Division |  |  |  |  |  |
| Barishal | 57 (10.2) | 437 (78.0) | 66 (11.8) | 275.01 | <0.000 |
| Chattagram | 181 (10.9) | 1215 973.5) | 257(15.5) |  |  |
| Dhaka | 250 (11.2) | 1847 (82.7) | 136 (6.1) |  |  |
| Khulna | 110 (10.6) | 893 (86.1) | 34 (3.3) |  |  |
| Mymensingh | 77 (11.0) | 507 (72.4) | 116 (16.6) |  |  |
| Rajshahi | 138 (11.8) | 973 (83.1) | 60 (5.1) |  |  |
| Rangpur | 107 (10.5) | 828 (81.3) | 84 (8.2) |  |  |
| Sylhet | 61 (12.2) | 344 (68.9) | 94 (18.8) |  |  |

| Residence |  |  |  |  |  |
| --- | --- | --- | --- | --- | --- |
| Urban | 293 (11.6) | 2069 (82.2) | 155 (6.2) | 47.08 | <0.000 |
| Rural | 689 (10.8) | 4973 (78.3) | 693 (10.9) |  |  |
| Religion |  |  |  |  |  |
| Muslim | 907 (11.3) | 6310 (78.7) | 805 (10.0) | 29.58 | <0.000 |
| Non-Muslim | 75 (8.8) | 732 (86.1) | 43 (5.1) |  |  |
| Wealth index |  |  |  |  |  |
| Poorest | 140 (9.0) | 1171 (75.5) | 241 (15.5) | 138.04 | <0.000 |
| Poorer | 179 (10.4) | 1348 (78.1) | 198 (11.5) |  |  |
| Middle | 202 (10.8) | 1492 (79.6) | 180 (9.6) |  |  |
| Richer | 224 (12.1) | 1478 (80.2) | 142 (7.7) |  |  |
| Richest | 238 (12.7) | 1553 (82.7) | 87 (4.6) |  |  |
| Mother's Education |  |  |  |  |  |
| No | 40 (3.8) | 719 (87.8) | 302 (28.5) | 902.09 | <0.000 |
| Primary | 112 (5.1) | 1755 (80.1) | 324 (14.8) |  |  |
| Secondary | 579 (13.5) | 3505 (81.6) | 212 (4.9) |  |  |
| Higher | 251 (19.0) | 1062 (80.3) | 10 (0.8) |  |  |
| Father's Education |  |  |  |  |  |
| No | 74 (4.1) | 1359 (74.5) | 392 (21.5) | 670.91 | <0.000 |
| Primary | 198 (7.9) | 2054 (82.0) | 254 (10.1) |  |  |
| Secondary | 370 (12.7) | 2379 (81.6) | 165 (5.7) |  |  |
| Higher | 339 (20.8) | 1251 (76.9) | 37 (2.3) |  |  |

| Father's Occupation |  |  |  |  |  |
| --- | --- | --- | --- | --- | --- |
| Did not work | 38 (13.4) | 181 (64.0) | 64 (22.6) | 279.41 | <0.000 |
| Agriculture | 145 (6.8) | 1643 (77.0) | 345 (16.2) |  |  |
| Worker/labor | 678 (12.2) | 4458 (80.5) | 404 (7.3) |  |  |
| Professional | 115 (12.8) | 750 (83.5) | 33 (3.7) |  |  |
| Business & others | 6 (35.3) | 10 (58.8) | 1 (5.9) |  |  |
| Mother's Occupation |  |  |  |  |  |
| Did not work | 780 (13.9) | 4305 (76.9) | 510 (9.1) | 202.92 | <0.000 |
| Agriculture | 73 (3.8) | 1611 (83.1) | 255 (13.2) |  |  |
| Worker/labor | 93 (9.8) | 798 (83.7) | 62 (6.5) |  |  |
| Professional | 20 (9.7) | 180 (87.0) | 7 (3.4) |  |  |
| Business | 8 (9.5) | 74 (77.9) | 13 (13.7) |  |  |
| Others | 8 (9.5) | 75 (89.3) | 1 (1.2) |  |  |
| Types of toilet facilities |  |  |  |  |  |
| Toilet with Flush | 332 (11.2) | 2419 (81.9) | 202 (6.8) | 278.17 | <0.000 |
| VIP Latrine | 162 (8.8) | 1502 (81.4) | 181 (9.8) |  |  |
| Pit Latrine | 286 (8.7) | 2571 (78.2) | 431 (13.1) |  |  |
| Hanging toilet and others | 201 (25.6) | 550 (70.2) | 33 (4.2) |  |  |
| Water Sources |  |  |  |  |  |
| Other than<br>Tubewell or<br>Borehole | 324 (16.7) | 1526 (78.6) | 92 (4.7) | 131.47 | <0.000 |
| Tubewell or<br>Borehole | 658 (9.5) | 5516 (79.6) | 756 (10.9) |  |  |
| Household Member |  |  |  |  |  |
| 1 to 3 Members | 301 (14.8) | 1648 (81.3) | 78 (3.8) | 199.86 | <0.000 |
| 4 to 6 Members | 466 (9.0) | 4205(81.1) | 512 (9.8) |  |  |
| More than 6<br>Members | 215 (12.9) | 1190 (71.5) | 259 (15.6) |  |  |
| BMI Category |  |  |  |  |  |
| Thin | 144 (16.6) | 634 (73.1) | 89 (10.3) | 74.92 | <0.000 |
| Normal | 575 (12.1) | 3691 (77.9) | 473 (10.0) |  |  |
| Overweight | 197 (7.7) | 2140 (83.9) | 213 (8.4) |  |  |
| Obese | 66 (9.2) | 578 (80.5) | 74 (10.3) |  |  |
| Age at first marriage |  |  |  |  |  |
| Below 18 years | 499 (8.4) | 4724 (79.9) | 689 (11.7) | 192.66 | <0.000 |
| 18 years and<br>above | 483 (16.3) | 2318 (78.3) | 159 (5.4) |  |  |
| Contraceptive Use |  |  |  |  |  |
| Modern method | 217 (4.70) | 3947 (86.3) | 411 (9.0) | 616.27 | <0.000 |
| Traditional<br>methods | 44 (5.0) | 701 (78.9) | 143 (16.1) |  |  |
| No methods | 721 (21.1) | 2394 (70.2) | 294 (8.6) |  |  |
| **Child died** |  |  |  | 162.12 | <0.000 |
| No | 946 (12.0) | 6287 (79.7) | 660 (8.4) |  |  |
| Yes | 982 (11.1) | 755 (77.1) | 188 (19.2) |  |  |
| **Access to media** |  |  |  |  |  |
| No | 370 (10.0) | 2798 (75.5) | 539 914.5) | 184.17 | <0.000 |
| Yes | 611 (11.8) | 4144 (82.2) | 309 (6.0) |  |  |
| **Sex of Household head** |  |  |  |  |  |
| Male | 850 (11.0) | 6099 (79.2) | 754 (9.8) | 3.58 | >0.166 |
| Female | 132 (11.3) | 943 (80.7) | 94 (8.0) |  |  |
| **Ideal number of children** |  |  |  |  |  |
| 0-2 Children | 819 (12.3) | 5446 (81.8) | 390 (5.9) | 575.53 | <0.000 |
| 3-4 Children | 154 (7.2) | 1577 (73.5) | 416 (19.4) |  |  |
| Over 4 Children | 9 (12.9) | 19 (27.1) | 42 (60.0) |  |  |
| **Husbands desire for children** |  |  |  |  |  |
| Both want same | 852 (11.00) | 6197 (79.9) | 706 (9.1) | 123.24 | <0.000 |
| Husbands wants more | 42 (7.4) | 439 (77.6) | 85 (15.0) |  |  |
| Husbands wants fewer | 21 (6.3) | 279 (84.3) | 31 (9.4) |  |  |
| Don't know | 67 (30.5) | 127 (57.7) | 26 (11.8) |  |  |
| **Father's desire for children** |  |  |  |  |  |
| One Children | 476 (39.7) | 719 (59.9) | 5 (0.4) | 2122.09 | <0.000 |
| Two Children | 360 (20.6) | 1385 (79.2) | 3 (0.2) |  |  |
| Three Children | 96 (14.6) | 552 (84.1) | 8 (1.2) |  |  |
| Four Children | 50 (0.9) | 4386 (83.3) | 832 (15.8) |  |  |

#### Data pre-processing

Data cleaning and pre-processing were carried out before statistical modeling to ensure analytical correctness and consistency. Observations with missing values in important variables were removed using list-wise deletion, yielding a final dataset with complete cases for all variables included in the analysis. All categorical variables were encoded using label encoding, which transformed textual categories into numerical labels appropriate for modeling. The cleaned dataset was then divided into training and testing subsets via an 80:20 train-test split. Specifically, 80% of the data was randomly assigned to the training set for model building, with the remaining 20% put aside for model validation.

#### Feature selection

A hybrid strategy for feature selection was applied, integrating Chi-square testing, Complementary Information, and Random Forest significance to find key predictors of pregnant adolescents. Every feature was evaluated according to its score in the three techniques, and an average ranking was calculated. Variables with the highest average rankings were chosen, guaranteeing strong and reliable selection across both statistical and model-based methods.

#### Machine learning model development

A thorough array of machine learning models was utilized to examine and forecast fertility trends among women in Bangladesh. These comprised Random Forest (RF), Decision Tree (DT), K-Nearest Neighbors (KNN), Logistic Regression (LR), Support Vector Machine (SVM), XGBoost, LightGBM, and Neural Network (NN), Stacking, Voting [1]. Along with individual classifiers, ensemble learning techniques like stacking and voting models were utilized to enhance predictive performance [30, 31].

### Model Evaluation and Validation

Performance was evaluated using a standard set of classification metrics, including accuracy, precision (weighted), recall (weighted), F1-score (weighted), and AUC-ROC (weighted). To measure the robustness and generalizability of each of these performances, metrics were calculated for various fold sizes using K-fold cross-validation (K = 5, 10, 15, 20, 30). In each fold iteration run, along with validation of the model on the fold, other K-1 folds were used for performing training. Performance is recorded for each fold, and the averaged performance is computed based on the K values. This reduces any possible influence of overfitting or selection bias while providing a reasonable estimate of model quality. Considering several K values allows observing the bias-variance trade-off, making it easier to select the best fold size for model stability.

### Software and Computational Tools

All analysis was conducted in Python (version 3.12) with the relevant libraries such as scikit-learn, pandas, and numpy for model estimation and training. SPSS (version 25) was also used for preliminary statistical analysis. A fixed random seed (42) was used for reproducibility of results.

### Ethics Statement

The study was based on publicly available secondary data from the Bangladesh Demographic and Health Survey (BDHS), which is collected under the authorization of the National Institute of Population Research and Training (NIPORT), Bangladesh, with technical support from ICF International. The BDHS survey protocols were reviewed and approved by the ICF Institutional Review Board (IRB) and the National Research Ethics Committee of Bangladesh.

All DHS datasets are fully anonymized before release, and no identifiable participant information is included. Therefore, this analysis did not require additional ethical approval. Permission to use the data was obtained through the DHS Program data request system.

## Results

**Table 1** illustrates the frequency distribution of the number of children ever born to mothers across various demographic and background characteristics, along with corresponding chi-square values and p-values indicating statistical significance. According to the table, the Dhaka division represents the highest proportion of mothers with 82.7% having 1 to 3 children, while Sylhet shows a notable fertility pattern with 18.8% having 4 or more children, the highest among all divisions. The Khulna division demonstrates the lowest percentage (3.3%) of mothers with 4 or more children and a high concentration (86.1%) within the 1 to 3 children category. In urban areas (11.6%), the majority of mothers (82.2%) have 1 to 3 children, with only 6.2% having 4 or more. In contrast, rural areas (689 mothers or 10.8%) show a slightly lower percentage in the 1 to 3 children category (78.3%) and a higher proportion (10.9%) with 4 or more children, indicating a rural-urban disparity in fertility rates. The data by religion reveals that Muslim mothers make up the majority (11.3%) with no children, 78.7% with 1 to 3 children, and 10% with 4 or more). Meanwhile, non-Muslims, although a smaller proportion of the population, display a lower tendency for higher fertility only 5.1% have 4 or more children, and 86.1% fall in the 1 to 3 children group. A closer look at the wealth index shows a clear gradient in fertility patterns. Poorest mothers have the highest percentage (15.5%) with 4 or more children and the lowest (9.0%) with no children, suggesting higher fertility among economically disadvantaged groups. In contrast, mothers in the richest wealth quintile have the lowest percentage (4.6%) with 4 or more children and the highest percentage (12.7%) with no children, while a significant majority (82.7%) have 1 to 3 children. This highlights a strong inverse relationship between wealth and high fertility. All these characteristics division (χ²=275.01, p<0.001), place of residence (χ²=47.08, p<0.001), religion (χ²=29.58, p<0.001), and wealth index (χ²=138.04, p<0.001)—show statistically significant associations with the number of children ever born, confirming that fertility patterns are influenced by geographical, socioeconomic, and cultural factors.

Moreover, the data demonstrates that both maternal and paternal education levels play a critical role in determining the number of children ever born. Among mothers with no education, 28.5% have 4 or more children, which is the highest fertility across all education groups, while only 3.8% have no children. In contrast, mothers with higher education exhibit significantly lower fertility only 0.8% have 4 or more children, while 19.0% remain childless, suggesting an inverse relationship between maternal education and fertility. Similarly, mothers with secondary education show a strong concentration (81.6%) in the 1 to 3 children category. Father’s education mirrors this trend. Fathers with no education have the highest proportion (21.5%) of wives with 4 or more children, while those with higher education show a much lower fertility rate only 2.3% have 4 or more children, and 20.8% have no children. The chi-square value of 670.91 (p<0.001) confirms the strong significance of paternal education on fertility outcomes. Father’s occupation also influences fertility outcomes. Fathers involved in agriculture or who did not work are associated with higher fertility (16.2% and 22.6% with 4 or more children, respectively). Meanwhile, professional workers and those in business/other categories have lower fertility, with only 3.7% and 5.9%, respectively, having 4 or more children. Notably, professional workers show the highest concentration (83.5%) in the 1 to 3 children group. Mother’s occupation demonstrates varied fertility patterns. Mothers working in agriculture report 13.2% with 4 or more children, whereas professional women show only 3.4%, highlighting the correlation between occupational status and family size. Interestingly, housewives, the largest group, have 13.9% with no children and 9.1% with 4 or more, showing a mid-range fertility profile. Toilet facilities are also found to influence fertility. Households with flush toilets are more likely to have fewer children, with only 6.8% having 4 or more. In contrast, those using pit latrines or VIP latrines show increased fertility (13.1% and 9.8%, respectively). The highest rate of childlessness (25.6%) is observed among households with hanging toilets and other types, though most in this group (70.2%) have 1 to 3 children. Access to safe water sources plays a similarly important role. Households relying on tubewells or boreholes have 10.9% with 4 or more children and 9.5% with no children. In contrast, those using other water sources show lower fertility (4.7% with 4+ children) but a much higher percentage (16.7%) with no children, suggesting potential links between water insecurity and reproductive behavior.

Furthermore, household composition shows a clear pattern in fertility. Mothers in households with more than 6 members report the highest proportion (15.6%) with 4 or more children. Conversely, small households (1 to 3 members) report the lowest fertility (3.8%) and the highest proportion of mothers with no children (14.8%). This indicates that larger families may be associated with higher fertility, a relationship confirmed by a significant chi-square value (χ²=199.86, p<0.001).Nutritional status, reflected through BMI, also affects fertility. Thin women show the highest rate (16.6%) of childlessness, while those with a normal BMI cluster within the 1 to 3 children range (77.9%). Obese and overweight categories exhibit moderate fertility patterns, with 10.3% and 8.4% having 4 or more children, respectively. BMI status has a significant association with fertility (χ²=74.92, p<0.001), suggesting that maternal health may influence reproductive outcomes. Age at first marriage has a profound effect on fertility. Mothers who married before 18 show significantly higher fertility, with 11.7% having 4 or more children. Those who married at 18 or older have more childlessness (16.3%) and lower high-parity births (5.4%), indicating that early marriage contributes to higher fertility (χ²=192.66, p<0.001). The type of contraceptive method used has a strong association with fertility. Mothers using modern methods have lower fertility, with only 9.0% having 4 or more children and the majority (86.3%) with 1 to 3 children. Those using traditional methods show a much higher fertility rate (16.1%). Notably, women using no contraceptive method have the highest percentage of childlessness (21.1%), yet still maintain a significant portion with higher fertility (8.6%). These trends are statistically significant (χ²=616.27, p<0.001). The death of a child is related to increased fertility. Among these mothers, 19.2% had four or more children, compared to 8.4% who have never lost a child. This compensating behavior is significant (χ²=162.12, p<0.001) and indicates replacement-level fertility. Access to media is linked to lower fertility. Mothers who have media access are more likely to have one to three children (82.2%) and less likely to have four or more (6.0%).In comparison, those without access have a greater percentage (14.5%) with 4 or more children and lower rates of small family size, suggesting that media exposure may assist informed reproductive decisions (χ²=184.17, p<0.001). The sex of the household head has no statistically significant effect on fertility (χ²=3.58, p=0.166). Fertility patterns are generally comparable in male-headed households (9.8%) with four or more children and female-headed households (8.0%).Desired family size is closely aligned with actual fertility outcomes. Respondents favoring 0–2 children show low high-parity births (5.9%), while those preferring 3–4 children have a much higher rate (19.4%) with 4 or more children. The most striking result is among those who believe in having over 4 children, with 60.0% having 4+ children, confirming a strong alignment between fertility preferences and outcomes (χ²=575.53, p<0.001). Spousal preferences also affect fertility. When both partners agree, 9.1 % of mothers have four or more children. However, if the spouse wishes to have more children, the percentage increases to 15.0%, demonstrating that male fertility preferences can impact reproductive decisions. Interestingly, when the wife is confused about the husband’s wishes, childlessness rises significantly (30.5%), indicating potential communication or decision-making gaps (χ²=123.24, p<0.001).When both spouses consent, 9.1% of mothers have four or more children. However, if the husband chooses to have additional children, the number rises to 15.0%, suggesting that male fertility preferences might influence reproductive decisions. Interestingly, when the wife is unsure about the husband’s preferences, the number of children rises significantly (30.5%), indicating possible communication or decision-making gaps (χ²=123.24, p<0.001).All these characteristics division (χ²=275.01, p<0.001), place of residence (χ²=47.08, p<0.001), religion (χ²=29.58, p<0.001), and wealth index (χ²=138.04, p<0.001), mother’s education (χ²=902.09, p<0.001), father’s education (χ²=670.91, p<0.001), father’s occupation (χ²=279.41, p<0.001), mother’s occupation (χ²=202.92, p<0.001).

**Table 2** displays the performance of various machine learning models based on five essential metrics: accuracy, F1-score, precision, recall, and AUC ROC (all weighted). Logistic Regression (LR), LightGBM, Stacking, and Voting models performed best, with accuracy of 81%, F1-scores of 78%, precision values ranging from 78-79%, recall of 81%, and AUC scores of 81% (Fig 2). Random Forest (RF) and Neural Network (NN) both produced competitive results, with accuracy of 80-81% and F1-scores of 77%, while XGBoost followed closely with 79% accuracy and 77% F1-score. KNN performed moderately well, achieving 79% accuracy and 76% F1-score. In contrast, Decision Tree (DT) underperformed, particularly in AUC ROC (60%), indicating weak discriminative capacity. Notably, Support Vector Machine (SVM) exhibited an unusually low F1-score of only 20%, despite having high values for precision (80%) and recall (80%), possibly indicating severe class imbalance or skewed predictions. Overall, Logistic regression, LightGBM, Stacking, and Voting emerge as the most reliable models for predicting fertility-related factors, consistently above 80% across multiple evaluation parameters.

**Fig 2:**
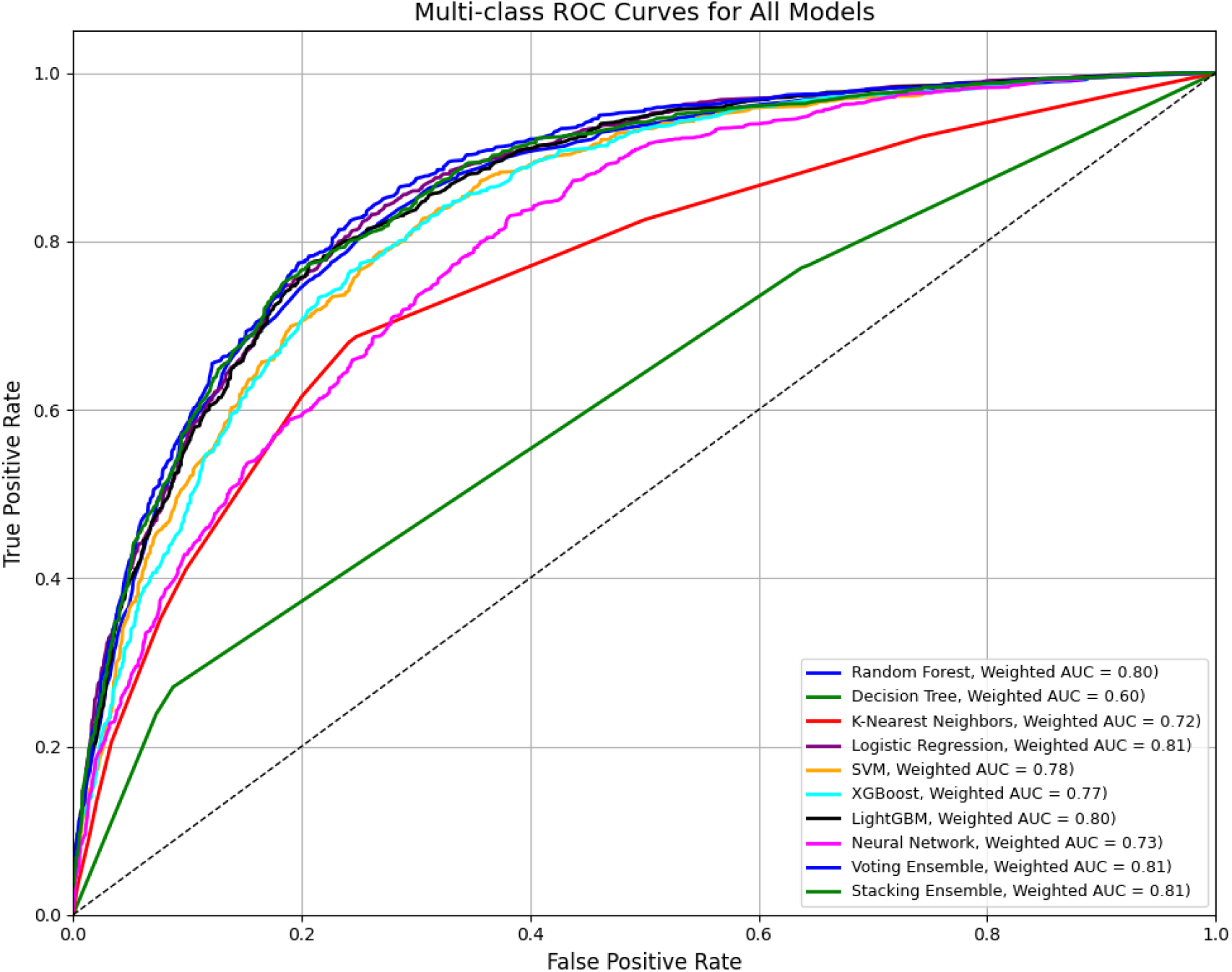
AUC–ROC curve for overall machine learning models

**Table 2:** Evaluation of different ML models performance.

| Model | Accuracy | F1-Score<br>(Weighted) | Precision<br>(Weighted) | Recall<br>(Weighted) | AUC ROC<br>(Weighted) |
| --- | --- | --- | --- | --- | --- |
| RF | 0.81 | 0.77 | 0.78 | 0.81 | 0.80 |
| DT | 0.72 | 0.72 | 0.73 | 0.72 | 0,60 |
| KNN | 0.79 | 0.76 | 0.76 | 0.79 | 0,72 |
| LR | 0.81 | 0.78 | 0.79 | 0.81 | 0.81 |
| SVM | 0.80 | 0.2 | 0.80 | 0.80 | 0.78 |
| XGBoost | 0.79 | 0.77 | 0.76 | 0.79 | 0.77 |
| LightGBM | 0.81 | 0.78 | 0.78 | 0.81 | 0.80 |
| NN | 0.80 | 0.77 | 0.77 | 0.80 | 0.73 |
| Staking | 0.81 | 0.78 | 0.78 | 0.81 | 0.81 |
| Voting | 0.81 | 0.77 | 0.78 | 0.81 | 0.81 |

**Table 3** displays the mean accuracy of multiple ML models using different K-Fold cross-validation setups (5 to 30 folds). Logistic Regression (LR), Voting, and LightGBM consistently achieved the best accuracies, all above 80% throughout multiple folds, with Voting peaking at 80.99% after 30 folds. Random Forest, XGBoost, and Stacking all achieved stable performance of around 80%. In contrast, Decision Tree (DT) achieved the lowest accuracy (71-72%), while SVM, KNN, and Neural Network produced moderate but consistent results. Overall, LR, Voting, and LightGBM demonstrated the best generalization performance across all fold variations.

**Table 3:** K-Fold cross validation of ML Model.

| Models | Mean Accuracy of K-Fold |  |  |  |  |
| --- | --- | --- | --- | --- | --- |
|  | 5-Fold | 10-Fold | 15-Fold | 20-Fold | 30-Fold |
| RF | 0.8016 | 0.8037 | 0.8036 | 0.8038 | 0.8033 |
| DT | 0.7172 | 0.7241 | 0.7258 | 0.7238 | 0.7235 |
| KNN | 0.7861 | 0.7937 | 0.7861 | 0.7915 | 0.7880 |
| LR | **0.8082** | 0.8085 | 0.8082 | **0.8084** | 0.8084 |
| SVM | 0.7957 | 0.7966 | 0.7975 | 0.7972 | 0.7975 |
| XGBoost | 0.7945 | 0.7960 | 0.7984 | 0.7981 | 0.8000 |
| LightGBM | 0.8040 | 0.8058 | **0.8088** | 0.8072 | 0.8055 |
| NN | 0.7796 | 0.7879 | 0.7878 | 0.7857 | 0.7861 |
| Stacking | 0.7987 | 0.7991 | 0.7983 | 0.8008 | 0.8009 |
| Voting | 0.8058 | **0.8087** | 0.8081 | 0.8083 | **0.8099** |

## Discussion

Overall, this research unveils the discussion on traditional methods and Machine Learning methods and we found that division, place of residence, religion, and wealth index, mother’s education, father’s education, father’s occupation, mother’s occupation, type of toilet facilities, and source of drinking water all show strong and statistically significant relationships with fertility levels, reinforcing the impact of education, employment, and infrastructure on reproductive behavior.

Moreover, water resources, Household Member, Body Mass Index, Age at first marriage, Contraceptive use, Child died, Access to Media, Ideal number of children, Husband desires for children, and Father’s desire for children were the significant factors for predicting fertility in Bangladesh. Our result provide similar prediction regarding significant factors with another study [1] conducted based on fertility determinants on 2024. Conflicts noticed in another important finding from a study is women who had a history of caesarean delivery were less likely to have high-risk fertility behavior. There are some other studies related to the association between type of delivery and subsequent fertility [34, 35]. Moreover, religious belief also did affect fertility. Our study revealed that the women who were Muslims were significantly associated with Number of child ever born compared with other religious believers. This finding was in line with an Indian study [36].

Our study presents the predictive ML model performance evaluation of various using metrics such as Accuracy, F1-Score, Precision, Recall, and AUC ROC, all expressed as percentages. Logistic Regression (LR), Random Forest (RF), and LightGBM, Staking, and Voting achieved the highest accuracy at 81%. Another study have found Support vector machine provide better prediction with 96.21% accuracy identified seven factors i.e., access to mass media, education level of father, mother educational level, number of household members, Body Mass Index (BMI), number of living children and incidents of sons or daughters died as most important as fertility determinant of Bangladesh[1].

Our research supports many of the arguments in the literature [37] that emphasize the strong relationship between fertility and Body Mass Index. We discovered that the likelihood of expressing a want to have no more children increases with the number of existing children, area of residence [28, 38].In our research found a strong association between fertility and number of household member, indicating that larger households tend to exhibit higher levels of fertility which is concurrent with the study [39].

## Conclusion

Machine Learning (ML) models is more reliable than traditional statistical models for determining fertility. The machine learning models use training data for constructing a good model and test data for predicting fertility in Bangladesh and then compare the reliability. With the help of our best identified model i.e., Logistic Regression (LR), Random Forest (RF), and LightGBM, Staking, and Voting achieved the highest accuracy at 81% for predicting the determinants of fertility in Bangladesh. However, LR, Staking, and Voting outperformed with AUROC value of 81% for predicting the determinants influencing the fertility in Bangladesh. In addition, All these characteristics division place of residence, religion, and wealth index, mother’s education, father’s education, father’s occupation, mother’s occupation, type of toilet facilities, and source of drinking water etc. all show strong and statistically significant relationships with fertility levels, reinforcing the impact of education, employment, and infrastructure on reproductive behavior. This study used the BDHS 2021-22 dataset and, being cross-sectional, cannot establish causality; self-reported data may be affected by memory errors or social desirability, and although eight machine learning models were used, future studies can explore additional models for better prediction.

## Data Availability

The data used in this study are publicly available from the Demographic and Health Surveys (DHS) Program. The Bangladesh Demographic and Health Survey (BDHS) 2022 dataset can be accessed upon registration and request through the DHS Program website: https://dhsprogram.com/data/. Interested researchers can obtain the data in the same manner as the authors. The authors had no special access privileges that others would not have.

